# The application of an extracellular vesicle-based biosensor in early diagnosis and prediction of chemoresponsiveness in ovarian cancer

**DOI:** 10.1101/2022.10.14.22281101

**Authors:** Meshach Asare-Werehene, Rob Hunter, Emma Gerber, Arkadiy Reunov, Isaiah Brine, Chia-Yu Chang, Chia-Ching Chang, Dar-Bin Shieh, Dylan Burger, Hanan Anis, Benjamin K. Tsang

## Abstract

Ovarian cancer (OVCA) is the most fatal gynecological cancer with late diagnosis and chemoresistance being the main obstacles of treatment success. Since there is no reliable approach to diagnosing patients at an early stage as well as predicting chemoresponsiveness, there is the urgent need to develop a diagnostic platform for such purposes. Extracellular vesicles (EVs) present as an attractive biomarker given their potential specificity and sensitivity to tumor sites. We have developed a novel sensor which utilizes cysteine functionalized gold nanoparticles to simultaneously bind to cisplatin (CDDP) and EVs affording us the advantage of predicting OVCA chemoresponsiveness, histologic subtypes, and early diagnosis using surface enhanced Raman spectroscopy. EVs were isolated and characterized from chemosensitive and resistant OVCA cells lines as well as pre-operative patient blood samples. The mechanistic role of plasma gelsolin (pGSN) in EV-mediated CDDP secretion in OVCA chemoresistance was investigated using standard cellular and molecular techniques. We determined that chemoresistant cells secrete significantly higher levels of small EVs (sEVs) and EVs containing CDDP (sEV-CDDP) compared with their sensitive counterparts. pGSN interacted with cortactin (CTTN) and both markers were significantly upregulated in chemoresistant patients’ tumors compared with the sensitive patients. Silencing pGSN decreased EV and EV-CDDP secretions in the resistant cells whereas its over-expression in sensitive cells upregulated EV and EV-CDDP secretion, suggesting the potential role of pGSN in EV-mediated CDDP export. sEV/CA125 ratio outperformed CA125 and sEV individually in predicting early stage, chemoresistance, residual disease, tumor recurrence, and patient survival. These findings highlight pGSN as a potential therapeutic target as well as providing a potential diagnostic platform to detect OVCA earlier and predict chemoresistance; an intervention that will positively impact patients’ survival.

## INTRODUCTION

Ovarian cancer (OVCA) remains the most fatal gynecological cancer with a 5-year survival rate of 45% primarily due to late diagnosis and chemoresistance (1-3). Although patients initially respond to treatment (primary surgical debulking and chemotherapy), about 50 – 70% of the patients relapse and become resistant to further treatment (3). The 5-year survival rate for stage 1 patients is >90% whereas that for stage 2, 3 and 4 are ∼70%, ∼39%, and ∼17% respectively (1, 3). However, only about 19% of ovarian cancer patients are diagnosed at the early stage despite the use of conventional biomarkers such as CA125, HE4, ROMA, and OVA1 (1-6). This suggests the urgent need for highly reliable and effective diagnostic platforms for early stage disease and chemoresistant prediction.

Plasma gelsolin (pGSN; also known as secreted GSN) is a multi-functional actin binding protein and the secreted isoform of the GSN gene (7-9). pGSN is implicated in the progression, chemoresistance, and metastasis of a plethora of cancer types including OVCA (10-18). We have previous demonstrated that pGSN is highly expressed in chemoresistant OVCA tumors and also transported via small extracellular vesicles (sEVs) (10, 15, 16). sEV-pGSN autoregulates its own gene expression and confers resistance to chemosensitive cells in a paracrine manner through the activation of the FAK/AKT/HIF1a pathway (10). Additionally, sEV-pGSN induces caspase-3 dependent apoptosis as well as suppresses the anti-tumor functions of immune cells such as CD8+ T cells, CD4+ T cells, and M1 macrophages (15, 16). Although pGSN is closely associated with sEV secretion, we have yet to determine the role of pGSN in sEV secretion.

EVs present as an attractive biomarker given they carry bioprints of the secreting cells and could capture the molecular landscape of cancer cells. This offers a superior advantage for cancer diagnostic tests compared with other biomarkers. EVs are secreted from most cells and are also present in liquid biopsies such as urine, plasma, and ascites. EVs are broadly categorized into apoptotic bodies (>1000 nm), large EVs (200 – 500 nm) and small EVs (30 – 300 nm) and serve as carriers for transporting molecular signatures such as proteins, RNA, DNA, and miRNAs (19, 20). The contents of the cargo could induce phenotypic and genetic changes in the recipient cells. sEVs play a key role in tumorigenesis and chemoresistance in different cancer types (15, 21-23). Chemoresistant OVCA cells secrete increased levels of sEVs containing pGSN which confers chemoresistance on otherwise chemosensitive cells as well as downregulate the tumor-killing functions of immune cells in the tumor microenvironment (10, 15, 16). Despite the potential diagnostic utility of sEVs, detecting EV specific antigens using conventional techniques like polymerase chain reaction (PCR) and enzyme-linked immunosorbent assay (ELISA) present as a major obstacle in cancer diagnostics. Thus, the use of technologies that could effectively detect the biochemical fingerprints of EVs will significantly enhance their clinical utility as opposed to targeting specific markers.

Surface enhanced Raman spectroscopy (SERS) has the capacity to examine the biochemical structure of biological analytes and presents as a useful tool in differentiating cancer cells from normal cells (24, 25). Interestingly, the application of SERS in liquid biopsy analyses has encountered major setbacks due to the heterogeneity of samples and interferences from large proteins. Similar obstacles are likely to be encountered in chemoresistance prediction given that not all circulating EVs originate from cancer cells, and those share biochemical characteristics with the target EVs. We have recently developed a novel sensor which utilizes cysteine functionalized gold nanoparticles to simultaneously bind to cisplatin (CDDP) and EVs affording us the advantage of quantifying EVs and EV-CDDP with high accuracy using SERS (26). This unique biosensor will enable us to overcome the challenges of tumor heterogeneity and protein interferences.

In this study, we investigated the application of an EV-based novel biosensor in the diagnosis of early-stage OVCA and the prediction of chemoresistance. Additionally, we examined the regulation of EV-mediated CDDP efflux by pGSN in chemoresistant OVCA cells. We found that pGSN is a key regulator of EV secretion and EV-mediated release of CDDP from chemoresistant OVCA cells. sEV/CA125 is also a strong indicator for stage 1 OVCA as well as predictor of chemoresistance.

## MATERIALS AND METHODS

### Ethics Statement

All the subjects recruited provided a written informed consent and the study was conducted in accordance with the appropriate guidelines approved by the Centre hospitalier de l’Université de Montreal (CHUM) Ethics Committee (IRB approval number; BD 04-002) and the Ottawa Health Science Network Research Ethics Board (IRB approval number; OHSN-REB 20150646-01H).

### Plasma Samples

Ninety-nine (99) ovarian cancer patients (high grade serous, HGS; 69, low grade serous, LGS; 4, not verified; 26) with predetermined CA125 levels and twenty (20) healthy non-cancerous subjects provided plasma samples for extracellular vesicle (EV) isolation, characterization, and analyses. Patients were recruited at the CHUM from 1992 – 2012 and did not receive any neoadjuvant chemotherapy or radiotherapy. All patients were managed with primary surgery. Gynecologic-oncologic pathologists examined all samples and assigned tumor grade and histologic subtypes in accordance with the International Federation of Gynecology and Obstetrics (FIGO) criteria. Computed Tomography imaging and CA125 levels during follow-ups were used to define disease-free survival (DFS; time of diagnosis to time of recurrence) and overall survival (OS; time of diagnosis to time of death). Details of patient demographics and clinical outcomes are outlined in **Supplementary Table S1**.

### Interrogation of OVCA public datasets

Ovarian cancer public datasets were interrogated using http://www.rocplot.org/ovarian/index and http://gepia.cancer-pku.cn/index.html on 06/16/2022. The differential expressions (box plot; chemoresistance vs. chemosensitive) and test performances (ROC curves; chemoresistance prediction) of GSN, CTTN, ABCB1, MRP2, and RAB27A (n=958; sensitive=862; resistant=96; platinum) were investigated. Spearman’s correlation tests were performed to assess the association between pGSN and the other genes. Significant correlations were inferred as *P* ≤ 0.05.

### Reagents

Cis-diaminedichloroplatinum (CDDP), phenylmethylsulfonyl fluoride (PMSF), aprotinin, dimethyl sulfoxide (DMSO), sodium orthovanadate (Na_3_VO_4_), CCK-8, and Hoechst 33258 were supplied by Millipore Sigma (St. Louis, MO). Two preparations of pGSN siRNA (siRNA1 and 2) and scrambled sequence siRNA (control) were purchased from Integrated DNA Technology (Iowa, USA) and Dharmacon (Colorado, USA), respectively. Human recombinant plasma gelsolin (hrpGSN) were synthesized and provided by Dr. Chia-Ching Chang, National Yang Ming Chiao Tung University, Taiwan. pGSN cDNA and 3.1A vector plasmids were provided by Dr. Dar-Bin Shieh, National Cheng Kung University Hospital, Taiwan. pCT-CD63-GFP was purchased from System Biosciences, LLC. See **Supplementary Table S2** for details on antibodies and other reagents.

### Ovarian Cancer Cell Lines

Chemosensitive and chemoresistant OVCA cell lines (1.6 × 10^6^ cells) of high grade serous (HGS; TOV3041G, TOV3133 and OV90) and endometrioid (A2780s and A2780cp) histologic subtypes were used for all *in vitro* studies. Dr. Anne-Marie Mes-Masson (CHUM, Montreal, Canada) generously donated the HGS cell lines whereas the endometioid cell lines were generously donated by Dr. Barbara Vanderhyden (Ottawa Hospital Research Institute, Ottawa, Canada). Quality control was performed to prevent any batch-to-batch changes on the morphology and growth rate. An addition, cell lines were regularly authenticated and tested for *Mycoplasma* contamination using PlasmoTest™ Mycoplasma Detection kit (InvivoGen; catalog number: rep-pt1). The HGS OVCA cells were maintained in OSE medium (wisent Inc. St-Bruno, QC, Canada) supplemented with 10% FBS (Millipore Sigma; St. Louis, MO, USA), 250 μg/mL amphotericin B, and 50 mg/mL gentamicin (wisent Inc. St-Bruno, QC, Canada). The endometrioid OVCA cells were maintained in Gibco RPM1 1640 (Life Technologies, Grand Island, NY, USA) supplemented with 10% FBS (Millipore Sigma; St. Louis, MO), 50 U/mL penicillin, 50 U/mL streptomycin, and 2 mmol/L l-glutamine (Gibco Life Technologies, NY, USA). All experiments were carried out in serum-free media. Details on the mutations of the cell lines used are described in **Supplementary Table S3**.

### pGSN Gene Interference

Chemoresistant cells were transfected with siRNA (50 nM, 24 h; scramble as controls) and sensitive cells transfected with cDNA (2 µg; 24 h; empty vectors as controls) using lipofectamine 2000, and were subsequently treated with CDDP (10 µM; 24 h)) then harvested for analysis as previously described (27-29). Two different siRNAs were used for each target to exclude off-target effects. Western blot was used to confirm pGSN knock-down and over-expression (30). See **Supplementary Table S2** for details on antibodies and **Supplementary Table S4** for customized siRNA oligonucleotide duplexes.

### Extracellular Vesicle (EVs) Isolation and Characterization

EVs were isolated and characterized from serum-free conditioned media from cultured cells as well as ovarian cancer patient plasma as described (31). Differential ultracentrifugation was used to isolate EVs from conditioned media while ExoQuick Ultra was used for the plasma samples. Conditioned media were centrifuged at 300 ×g (10 mins at RT) to remove cells and debris, 20,000 ×g (20 mins at RT) to remove large EVs (microparticles), and then at 100,000 ×g (90 mins at 4°C) to pellet sEVs. For plasma-derived EV isolation, 100 μL of ExoQuick precipitation reagent is added to plasma (40 μL in 500 μL of 0.1μm filtered PBS) and incubated on ice for 30 mins. The content is then centrifuged at 3,000 ×g for 10 mins and the suspension discarded. The pellet is then re-suspended in a buffer (provided by the manufacturer), transferred into pre-washed ExoQuick Ultra columns, and then centrifuged at 1,000 ×g for 30 secs. The receiving tube is then detached and EVs collected. EVs are then characterized using nanoparticle tracking analyses (particle size distribution and concentration), Western blot (EV-specific markers) and transmission electron microscopy (EV size and purity). Isolated EVs that were not used immediately were suspended in PBS and stored at -80 °C for subsequent analyses.

### Nanoparticle Tracking Analysis (NTA)

EVs diluted in PBS were analyzed, using the ZetaView PMX110 Multiple Parameter Particle Tracking Analyzer (Particle Metrix, Meerbusch, Germany) in size mode, and ZetaView software version 8.02.28, as previously described (31, 32). With 11 camera positions, EVs were captured at 21 °C.

### sEV-GFP tagging and uptake

Chemosensitive and chemoresistant OVCA cell lines (1.6 × 10^6^ cells) were transfected with exosome cyto-tracer, pCT-CD63-GFP (SBI System Biosciences; CYTO120-PA-1; 1 µg) in serum-free RPM1-1640 and then treated with CDDP (10 μM; 24 h). Cells were fixed and processed for confocal microscopy. Details on antibodies are described in **Supplementary Table S2**.

### Protein Extraction and Western blot Analysis

Western blotting (WB) procedure for proteins were carried out as described previously (27, 28, 30). After protein transfer, membranes were incubated with primary antibodies (1:1000) in 5% (wt/vol) blotto and subsequently with the appropriate horseradish peroxidase (HRP)-conjugated secondary antibody (1:2000) in 5% (wt/vol) blotto. See **Supplementary Table S2** for details of antibodies used. Chemiluminescent Kit (Amersham Biosciences) was used to visualize the peroxidase activity. Signal intensities generated on the film were measured densitometrically using Image J.

### Assessment of Cell Proliferation and Apoptosis

Apoptosis and cell proliferation were assessed morphologically with Hoechst 33258 nuclear stain and colorimetrically with the CCK-8 assay, respectively. “Blinded” counting approach was used to prevent experimental bias with the Hoechst 33258 nuclear staining.

### Transmission Electron Microscopy (TEM)

OVCA cell were pelleted (4000 ×g; 20 min) and processed, as previously described (33). Resin sections were stained with uranyl acetate and lead citrate solutions and examined with a Jeol JEM 1230 transmission electron microscope (Akishima, Japan).

### Immunoelectron Microscopy (iEM)

Cell pellets (4000 g; 20 min) were processed as previously described (33). The grids were washed three times in PBST, immunostained with anti-pGSN **antibody (Supplementary Table S2)**, rinsed in distilled water, stained with uranyl acetate and lead citrate, and photographed with a Jeol JEM 1230 transmission electron microscope (Akishima, Japan).

### EDS and ICP-MS

Energy Dispersive Spectroscopy (EDS) was performed in High-Resolution Transmission Electron Microscopy (HR-TEM, JEM-2010/JEOL Co. 200 KV) in NCKU for the quantitative analysis of elemental compositions and distributions in chemoresistant cells, particularly platinum signal that represent the anti-cancer drug. The samples were dispersed in ethanol and dropped onto a copper grid with an amorphous carbon film followed by evaporation of the solvent in a vacuum desiccator. The elemental composition was detected and quantified by EDS. In order to determine the Cisplatin content in the sEVs and in the condition medium, the Pt ion present in the drug was analyzed by THERMO Element XR High-resolution ICP-mass spectrometer (Element XR, Thermo Fisher Scientific, Bremen, Germany). Conditioned media were collected and sEVs were isolated using differential ultra-centrifugation before analysis. The sEVs pellets were lysed with lysis buffer (1% Triton X-100, 0,1 M Tris-HCl pH 7.4, 0.1% SDS) containing protease inhibitors (Sigma-Aldrich). Then, the sEVs lysates were digested by adding 200 µL or 50 µL of concentrated nitric acid (Optima Grade, Fisher Scientific, Cambridge, MA) and kept at at 60°C for 2 hours. The final digested solutions were diluted with deionized water (DI Water). Indium (1 µg/L) was added to the specimens as internal standard. The same matrix (lysing solution, nitric acid) was used as the calibration standard for the external control. Cell culture medium samples were diluted 1:100 with DI Water before performing the Pt analysis, adding only Indium as internal standard to minimize the effect of instrumental variation. The levels of the drug into the medium were expressed as ng of Cisplatin per mL of the solution.

### SERS quantification of sEV and CDDP

Cysteine capped gold nanoparticles were synthesized using the citrate reduction method to form particles from Au^3+^. After the particles had formed, they were washed via centrifugation and re-suspended in 10 mM NaOH with 1 µM cysteine. The amino acid preferentially bound to the gold surfaces due to the formation of a gold-thiol bond. To analyse sEV samples, they were first sonicated for 15 minutes to break-up the vesicles before mixing with the nanoparticles and incubating overnight. Before Raman measurement NaCl was added to the sEV-nanoparticle mixture to initiate aggregation, and this process was measured by means of an in-house built Raman spectrometer (785 nm excitation, 30 mW, 0.65 NA objective, Kaiser f/18i spectrograph, TE cooled Andor CCD). CDDP concentration could be inferred based on the aggregation rate of the particles, with increasing drug concentration resulting in faster aggregation (26). The steady-state SERS spectrum contained the signature of the sEV components which had also bound to the nanoparticles, and this spectrum could be used to quantify the sEVs in the sample using multi-variate regression (26).

### Statistical Analyses

The SPSS software version 25 (SPSS Inc., Chicago, IL, USA) and Graphpad Prism 7 (San Diego, CA, USA) were used to perform all statistical analyses and two-sided P ≤ 0.05 considered to indicate statistical significance. Receiving operating characteristic (ROC) curves were used to assess the performances of sEV/CA125, sEV and CA125 over their entire range of values. The area under the curve (AUC) was used as an index of global test performance. The association between sEVs and CA125 was analysed using Pearson’s correlation test (two-tailed). The means of sEVs and sEV/CA125 levels were plotted against stage, residual disease, histological subtypes, tumor recurrence, chemoresistance and survival using scatter plots and statistical analyses performed by using unpaired *t*-test; Gaussian distribution was tested. The relationship of these dichotomous variables to other clinicopathologic correlates was examined using Fisher exact test, T test and Kruskal Wallis Test as appropriate. Survival curves (DFS and OS) were plotted with Kaplan Meier and P-values calculated using the log-rank test. Univariate and multivariate Cox proportional hazard models were used to assess the hazard ratio (HR) for CA125, pGSN, stage (FIGO), residual disease and age as well as corresponding 95% confidence intervals (CIs).

## RESULTS

### Extracellular vesicle isolation and characterizations from OVCA cell lines

To investigate the role of pGSN in EV-mediated secretion of CDDP in OVCA chemoresistance, OVCA cell lines of HGS and Endometrioid subtypes were cultured with and without CDDP treatment (10 μM; 24 h) and their conditioned media were collected for EV isolation and characterization. Conditioned media was filtered with 0.22um pore size filter and EVs isolated using differential ultra-centrifugation (Fig. 1A). EVs were characterized using nanoparticle tracking analyses to determine particle size distribution and concentration (Fig. 1B), immune-gold electron microscopy to determine particle size, EV purity and gelsolin content (Fig. 1C). Based on the particle isolation and characterizations, the EVs had an average size of 130 nm and were positive for CD63 and CD9 markers suggesting they are sEVs. Electron microscopy confirmed that they contained secreted gelsolin (Fig. 1). SEVs from the plasma of OVCA patients and non-OVCA subjects were isolated using ExoQuick ULTRA (System Biosciences), which employs the use of a polymer for robust vesicle precipitation, low speed centrifugation, and column filtration (Fig. 1D). SEVs were characterized using nanoparticle tracking analysis and Western blotting (Fig. 1E). Unlike the sEV-free plasma, the sEVs isolated from patients’ plasma had an average median size of 136 nm and were positive for sEV markers (CD 9, CD63, and CD81) but negative for GM130, suggesting they are mainly sEVs (Fig. 1E and F). The presence of GAPDH also suggests the sEVs were intact (Fig. 1F).

**Figure 1:**
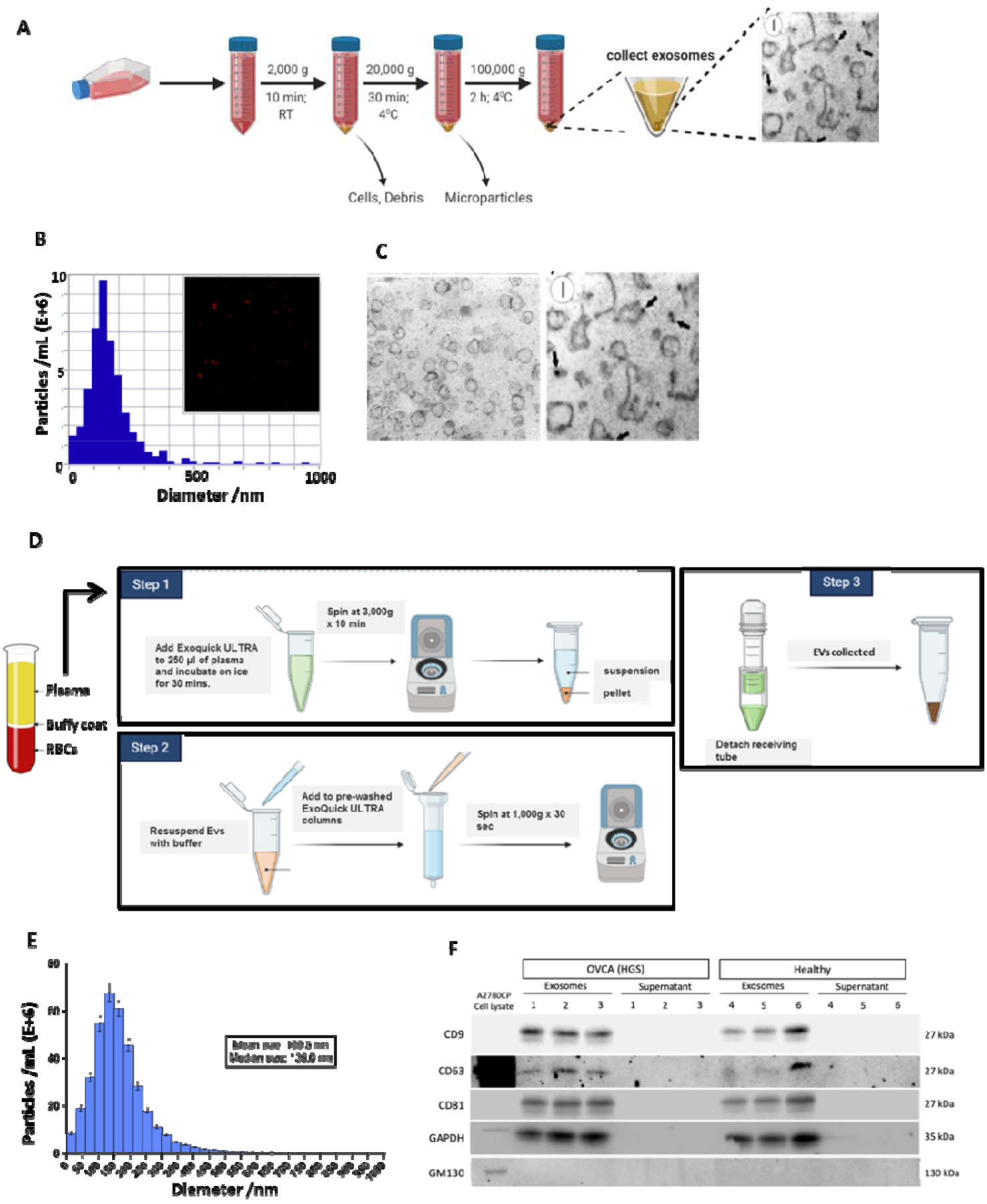
Extracellular vesicle (EVs) isolation and characterization. (A) Conditioned media form cultured OVCA cells were collected and filtered using 0.22 um filter. Differential ultra-centrifugation was used to isolate small EVs after which they were characterized using (B) nanoparticle tracking analyses (NTA; particle size distribution and concentration) and (C) electron microscopy (EM). (D) Small EVs were isolated from the plasma of human OVCA patients (n=99) and non-OVCA subjects (n=20) using Exoquick Ultra technique. EVs were then characterized using (E) nanoparticle tracking device (size distribution) and (F) Western blot analyses (small EV markers; CD9, CD63, CD81, negative marker; GM130, vesicle integrity marker; GAPDH).

### Chemoresistant OVCA cells secrete higher sEV-CDDP and exhibit a lateral EV secretion pattern

Chemosensitive (A2780s) and chemoresistant (A2780cp) cells were treated with or without CDDP (10 μM; 24 h) in a serum-free conditioned media (Fig. 2). The conditioned media was collected and sEVs isolated. CDDP concentrations were determined in the sEVs as well as EV-free conditioned media using inductively coupled plasma reactive ion etching (ICP) (Fig. 2A). We observed that sEVs derived from chemoresistant cells contain significantly higher levels of CDDP compared with that of chemosensitive cells (Fig. 2 A). Meanwhile, there were no significant differences in the CDDP concentration detected in the serum-free conditioned media of both cells (Fig. 2A). The OVCA cells collected were processed for transmission and immune-gold electron microscopy (Fig. 2B-C). We observed a lateral or peripheral secretion of EVs in more than 50% of the chemoresistant cells when treated with CDDP; a phenomenon that was not seen in chemosensitive cells as well as non-treated chemoresistant cells (Fig. 2B). Colloidal gold granules labeling pGSN were also present in EVs (Fig. 2B). Interestingly, the conventional EV secretion via multivesicular bodies and exocytosis were seen in chemoresistant cells not treated with CDDP as well as the chemosensitive cells (Fig. 2C). We also assessed the platinum distribution in the chemoresistant cells after CDDP treatment using energy dispersive x-ray spectroscopy (EDS) (Fig. 2D). Cellular distribution of platinum was not significantly different between chemoresistant cells treated with CDDP and those not treated (Fig. 2D), suggesting that CDDP might have been exported via sEV secretion.

**Figure 2:**
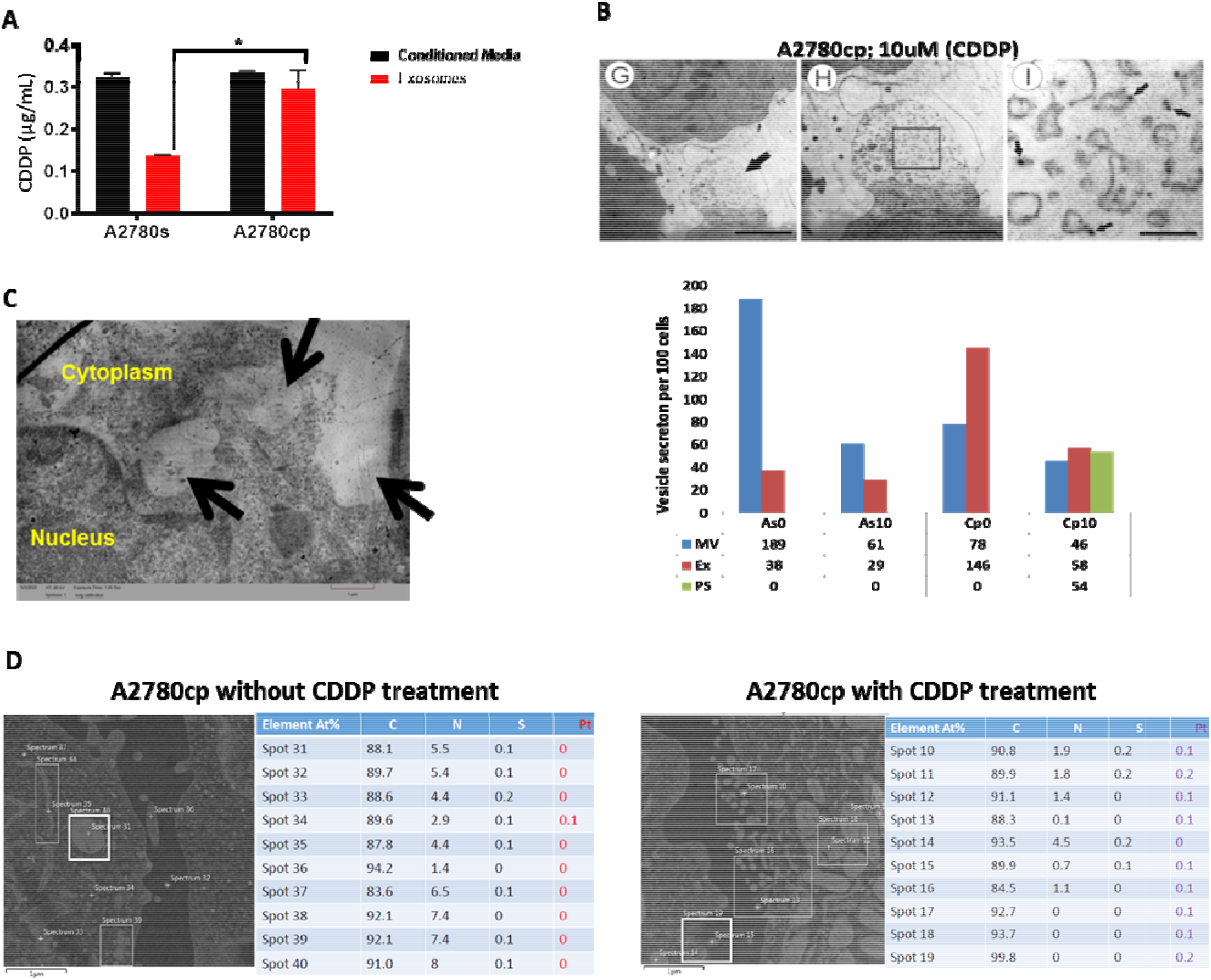
Chemoresistance OVCA cells secrete higher sEV-CDDP and exhibit abnormal EV secretion pattern. (A) sEV-CDDP content is higher in chemoresistant cells compared with chemosensitive cells. Chemosensitive (A2780s) and chemoresistant (A2780cp) were cultured and treated with or without CDDP (10 um; 24 h). Conditioned media were collected and sEVs isolated using differential ultra-centrifugation. CDDP content in the sEVs and conditioned media was analyzed using inductively coupled plasma reactive ion etching (ICP). (B and C) Chemoresistance OVCA cells when treated with CDDP exhibits peripheral secretion of sEVs which is not found in untreated chemoresistant cells. Chemoresistance OVCA cells were treated with or without CDDP (10 uM; 24 h). Cells were processed and sEV secretion analyzed using transmission and immuno-gold electron microscopy (As0; A2780s without treatment, As10; A2780s treated with 10 μM CDDP, Cp0; A2780cp without treatment and Cp10; A2780cp treated with 10 μM CDDP. (D) Platinum (Pt), calcium (C), nitrogen (N) and Sulphur (S) distribution within chemoresistant cells were analyzed using energy dispersive x-ray spectroscopy (EDS).

### pGSN and CTTN are highly expressed in human chemoresistant OVCA tumors and their protein contents in cell lines do not reduce upon CDDP treatment

sEV-mediated secretion of CDDP is a key determinant of OVCA chemoresistance; however, the underlying mechanism is not known. Given pGSN has been shown to be a marker of chemoresistance in OVCA as well as play a major role in EV biology; we investigated its potential role in sEV-mediated OVCA chemoresistance. We interrogated string-db.org and found that while there was no interaction between pGSN and drug transport proteins (p-glycoprotein; ABCB1 and ABCC2) as well as RAB27A, there was a potential interaction between pGSN and CTTN (Fig. 3A). We further interrogated the ovarian cancer TCGA public dataset (http://gepia.cancer-pku.cn/index.html) on 06/16/2022 to assess the correlation (Spearman coefficient) between pGSN and CTTN, RAB27A, or ABCB1. We found that pGSN positively correlated with CTTN and both proteins were significantly elevated in chemoresistant tumors compared with chemosensitive tumors (Fig. 3B and Supp. Fig. S1). A positive correlation was found between pGSN and MRP2, p-gp, and RAB27A (Supp. Fig. S1B-C). Additionally, we interrogated OV datasets (http://www.rocplot.org/ovarian/index) on 06/16/2022 to determine the differential expression (box plot; chemoresistance vs. chemosensitive) and test performance (receiver operating characteristic (ROC) analysis; chemoresistance prediction) of GSN, CTTN, ABCB1, MRP2, and RAB27A (Fig. 3B and Supp. Fig. S1) (n=958; sensitive=862; resistant=96). All patients had serous tumors, had received platin-based treatment, and response was based on progression free survival at 6 months. Using ROC analysis, we examined the test performance of pGSN (area under the curve (AUC)=0.56; p=0.03) and CTTN (AUC: 0.6; p<0.0001) and found both markers are significantly predictive of OVCA chemoresistance (Fig. 3C). We observed that P-glycoprotein but not MRP2 nor RAB27A are significantly predictive of chemoresistance in human OVCA tumors (Supp. Fig. S1A).

**Figure 3:**
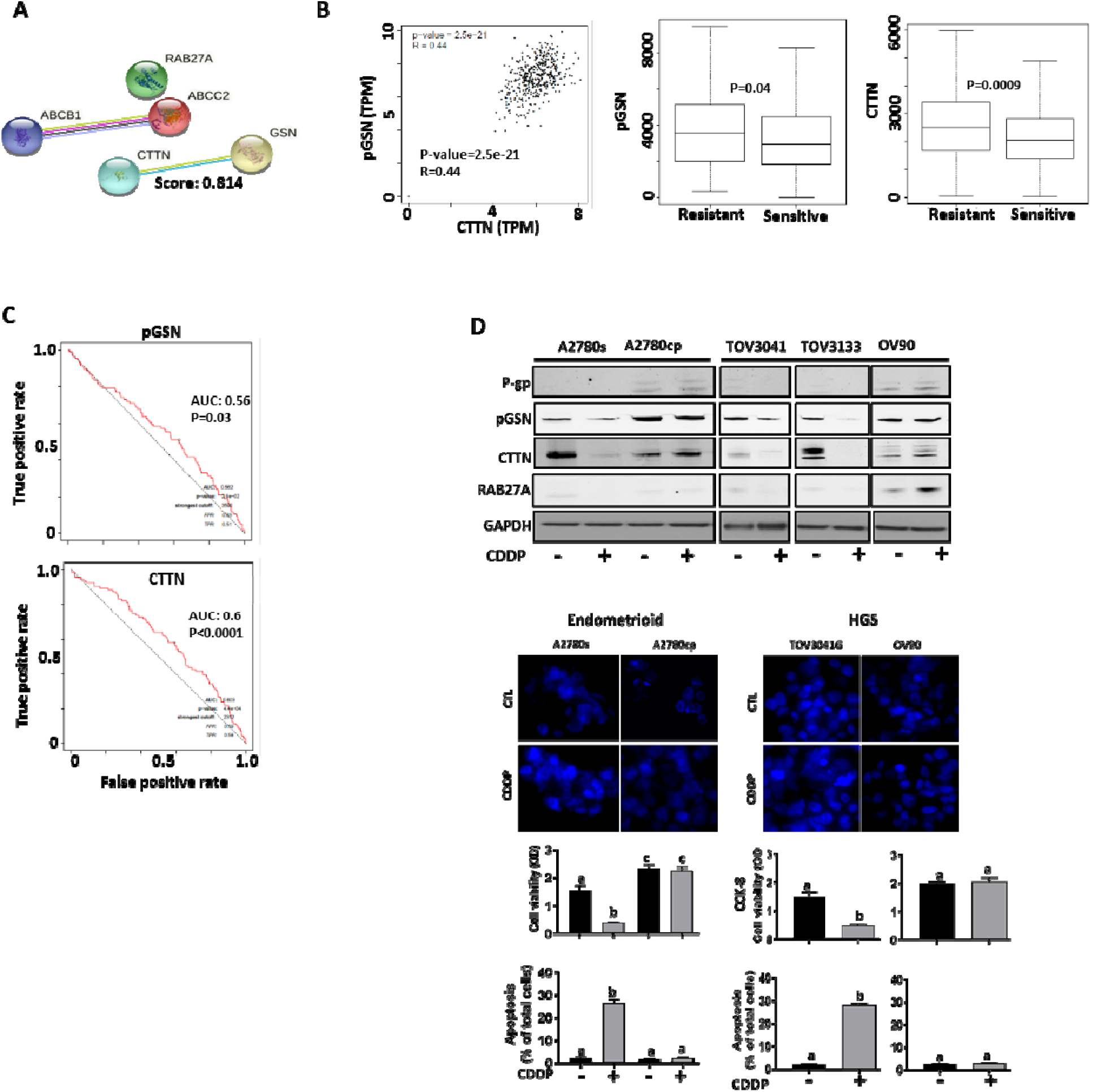
pGSN and CTTN are highly expressed in chemoresistant OVCA patient tumor and their protein contents in cell lines do not reduce upon CDDP treatment. (A) Using string-db.org, we found a strong interaction between pGSN and CTTN but not ABCB1 (p-gp), ABCC2, RAB27A. (B – C) Our investigation of TCGA public dataset revealed that pGSN positively correlates with CTTN expression and both proteins are highly elevated in chemoresistant OVCA patients (n=958). The test performances of pGSN and CTTN were evaluated using ROC curves and significant predictions were observed. (D) Chemosensitive (A2780s, TOV3041G, TOV3133) and chemoresistant (A2780cp and OV90) cells were treated with or without CDDP (10 uM; 24 h). pGSN, CTTN, P-gp, RAB27A and GAPDH contents were assessed by Western blot. Cisplatin-induced cell death was analysed by Hoechst staining and CCK-8 assay. Results are expressed as means ± SD from three independent replicate experiments. [D, (a; **P<0.01 vs b, a; ***P<0.001 vs c)].

To begin investigating the role of sEV-mediated chemoresistance in OVCA, chemosensitive (A2780s, TOV3041G, TOV3133) and chemoresistant (A2780cp and OV90) OVCA cells were treated with or without CDDP (10 μM; 24 h) (Fig. 3D). pGSN, CTTN, P-gp, RAB27A and GAPDH contents were assessed by Western blot. Cisplatin-induced cell death was analyzed by Hoechst staining, and cell viability was verified by CCK-8 assay (Fig. 3D). pGSN and CTTN contents were reduced by CDDP in the chemosensitive cells but not in the chemoresistant cells (Fig. 3D). These results were independent of histologic subtype. Notably, pGSN content was higher in chemoresistant compared to chemosensitive cells (Fig. 3D). P-glycoprotein and RAB27A contents were not detectable in the sensitive cells; however, weak signals were observed in the chemoresistant cells (Fig. 3D). Also, CDDP-induced apoptosis was significantly higher in the chemosensitive cells compared with the chemoresistant cells (Fig. 3D).

### pGSN regulates sEV release of CDDP and chemoresponsiveness in OVCA cells

The role of pGSN in sEV-mediated release of CDDP was further investigated in OVCA cells. Gold nanoparticles (AuNP) were synthesized and capped with cysteine (Au-cys) as previously demonstrated and characterized. Due to the strong negative charge of Au-cys particles, they form a mono-stable colloid in the absence of sEVs containing CDDP (Fig. 4A and B). Upon the addition of sEV containing CDDP, the cysteine residues are attacked by CDDP resulting in the reduction of the surface charges of the Au-NP (Fig. 4A and B). This results in Au-NP-sEV-CDDP aggregation which could be measured by Surface Enhance Raman Spectroscopy (SERS); a response that is proportional to the concentration of sEV and CDDP. Mono-stable colloid formation and aggregations of the particles were confirmed using transmission electron microscopy (Fig. 4B).

**Figure 4:**
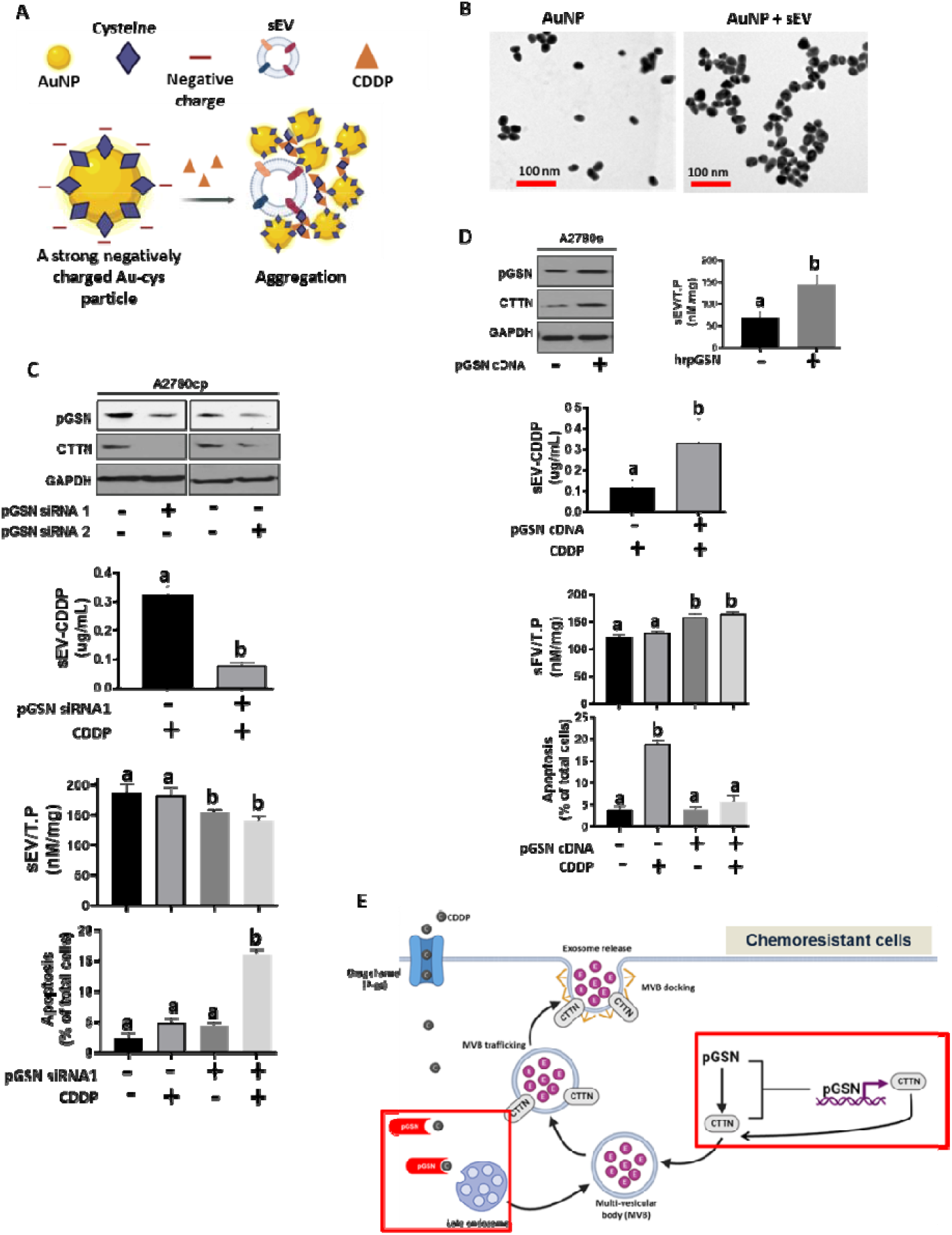
pGSN regulates sEV release of CDDP and chemoresponsiveness. pGSN downregulation in chemoresistant cells reduced CTTN content, sEV production and sEV-CDDP release whereas the vice-versa occurs when pGSN in over-expressed in chemosensitive cells. (A) Negatively charged cysteine capped gold particles (AuNP) react with CDDP and sEVs to form aggregates which is used for the SERS quantification. The rate of aggregation is proportional to the concentrations of CDDP and sEV. (B). Transmission electron microscope (TEM) images of mono-stable colloid of AuNP (without sEVs; left panel) and aggregated cysteine capped AuNP (with sEVs and CDDP; right panel). The biological components in the solution did not appear visible in the TEM images. pGSN was (C) silenced in chemoresistant cells (siRNA; 50 nM, 24 h) and (D) over-expressed in chemosensitive cells (cDNA; 2 ug, rhpGSN; 10 uM; 24 h). Cells were then treated with or without CDDP (10 uM; 24 h). pGSN, CTTN and GAPDH contents were assessed by Western blot and cisplatin-induced cell death analysed by Hoechst staining. sEV and CDDP concentrations were determined by biosensor aggregation using Raman spectroscopy. (E) A hypothetical model suggesting the role of pGSN in the sEV release of CDDP from chemoresistant OVCA cells. Results are expressed as means ± SD from three independent replicate experiments. [A - C, (a; **P<0.01 vs b)].

pGSN was silenced in chemoresistant cells using siRNAs (50 nM; 24 h; empty vector as a control) (Fig. 4C) and overexpressed in chemosensitive cells using cDNA (2 μg; 24 h) and human recombinant pGSN (hrpGSN, 10 μM; 24 h) (Fig. 4D and Supp. Fig. S2A). pGSN, CTTN and GAPDH contents were assessed by Western blot and CDDP-induced death analyzed by Hoechst staining. sEVs were isolated and their levels together with sEV-CDDP concentrations determined using SERS. pGSN knock-down resulted in the downregulation of CTTN content; a phenomenon that decreased sEV secretion as well as sEV-CDDP content (Fig. 4C and Supp. Fig. S2B). Additionally, pGSN knock-down sensitized the chemoresistant cells to CDDP-induced apoptosis (Fig. 4C). pGSN overexpression in chemosensitive cells increased CTTN content, sEV secretion as well as sEV-CDDP content (Fig. 4D and Supp. Fig. S2C). pGSN overexpression also suppressed CDDP-induced apoptosis in the chemosensitive cells (Fig. 4D). Additionally, we observed that hrpGSN promoted sEV secretion in chemosensitive cells; an observation which was consistent with pGSN overexpression by cDNA (Fig. 4D).

The above findings potentially suggest that pGSN regulates CTTN expression, which leads to enhanced secretion of sEVs in chemoresistant OVCA cells. Also, pGSN is enriched with sulphur and metallic binding sites; properties that might facilitate its direct binding to platinum, leading to enhanced CDDP packaging in sEVs and eventual release from the cell (Fig. 4E).

### SEV to CA125 ratio (sEV/CA125) is a strong predictor of stage 1 disease, chemoresponsiveness and favorable survival outcomes in OVCA patients

After characterizing our biosensor in OVCA cell lines, we extended its application to measuring SEV concentration in human patient plasma. Plasma samples were collected from 99 OVCA patients with HGS, LGS and unverified histologic subtypes **(Supplementary Table S2)**. Plasma-derived sEVs from OVCA patients and non-OVCA subjects were isolated and characterized (Fig. 1). The biosensor application was tested with the buffers used in the sEV isolation to ensure they do not interfere with the SERS measurement (Supp. Fig. S3A and B). Although the buffers increased the aggregation of the nanogold particles, this could easily be normalized and did not affect CDDP quantification (Supp. Fig. 3B and C). Mono-stable colloid formation and Au-NP aggregation with the plasma-derived sEV were confirmed using transmission electron microscopy before SERS quantification was performed (Fig. 5A).

**Figure 5:**
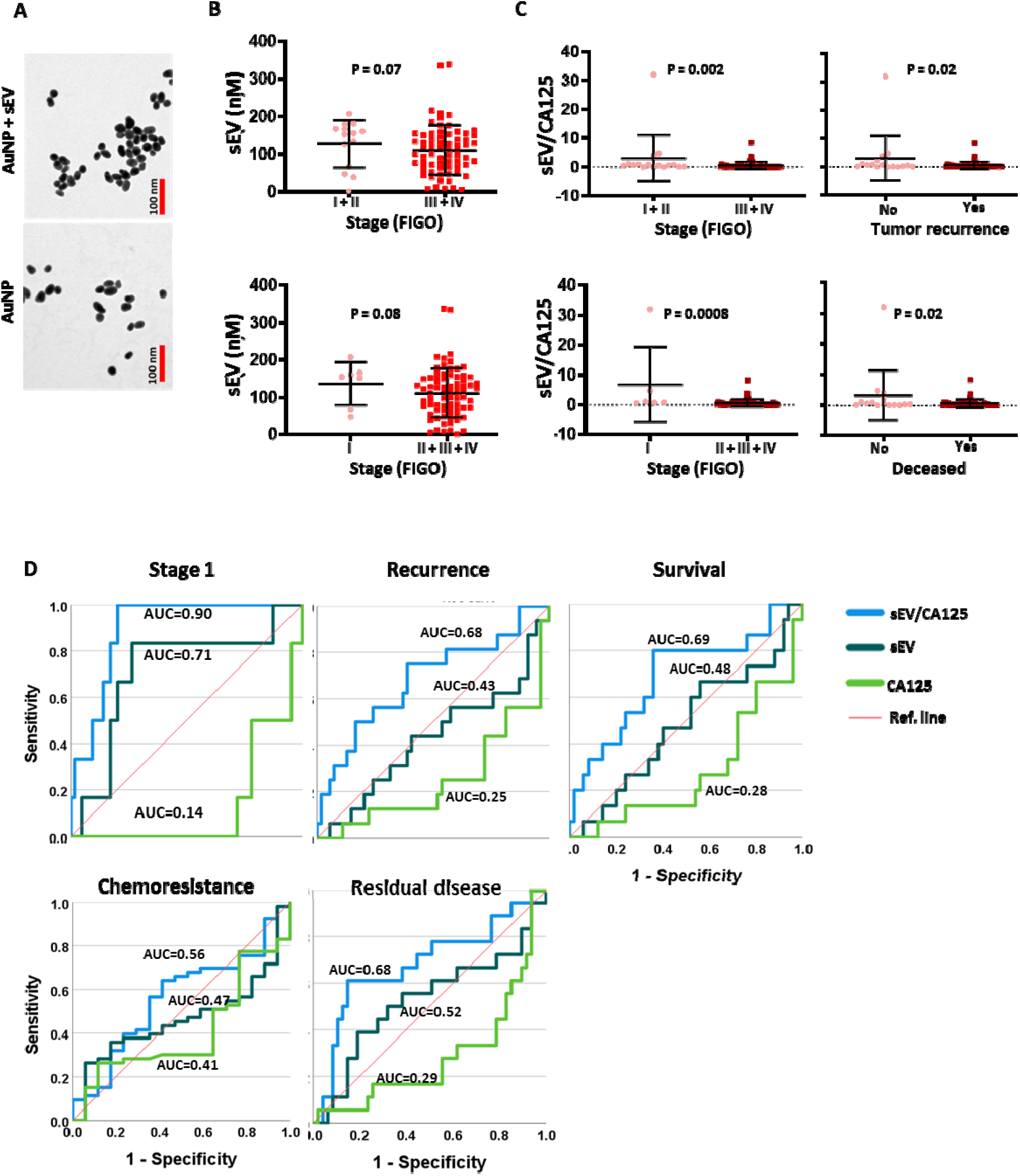
sEVCA125 predicts stage 1 and tumor recurrence with high test accuracy. (A) Transmission electron microscope (TEM) images of mono-stable colloid of AuNP (without plasma-derived sEVs; left panel) and aggregated cysteine capped AuNP (with plasma-derived sEVs; right panel). The biological components in the solution did not appear visible in the TEM images. SEVs were isolated from the plasma of human OVCA patient with pre-determined CA125, sEV or sEV/CA125 were correlated with patient clinical outcomes. (B) The mean levels of sEV concentrations were compared between early (I + II) and late (III + IV) stages as well as stage I and >stage 1 groups. Patients with late stage OVCA had lower levels of sEV concentration compared with early stage or stage 1 patients. (C) The mean levels of sEV/CA125 were correlated with OVCA clinical outcomes (stages, tumor recurrence and survival). (D) The test performances of sEV, sEV/CA125 and CA125 in the prediction of stage 1, tumor recurrence, survival, chemoresistance and residual disease were compared using ROC curves.

The sEV concentration and EV/CA125 ratio were determined and their mean±SD values correlated with patient clinical outcomes (stage, tumor recurrence, survival, chemoresistance, and residual disease) (Figs. 5, 6 and Supp. Fig. S4). We observed that patients with advanced stages of OVCA had levels of sEVs comparable to those in early stages (Fig. 5B); however, no significant difference in sEV concentrations was observed among the patients and the control healthy individuals. There’s also no correlation detected with the presence of residual disease (RD<1cm vs, RD>1cm), chemoresistance (PFI<6mo vs. PFI>6mo), tumor recurrence (yes vs. no), and the level of CA125 (low vs. high) (Supp. Fig. S4A). SEVs had a negative correlation with CA125, hence we developed a ratio with both markers to investigate their clinical utility (Supp. Fig. S4B). SEV/CA125 ratio had better clinical utility compared to sEV and CA125 concentrations alone with regards to OVCA stage, tumor recurrence and disease state (Fig. 5C). However, a trend (although not significant) was observed with residual disease and histologic differentiation subtypes (Supp. Fig. S4C). Using ROC analysis, we determined the test performances of sEV, sEV/CA125, and CA125 in predicting patient clinical outcomes (Fig. 5D). We found that sEV/CA125 was the most robust in predicting stage 1 disease, non-recurrence, patient survival, chemoresponsiveness and optimal residual disease in OVCA patients (Fig. 5D).

**Figure 6:**
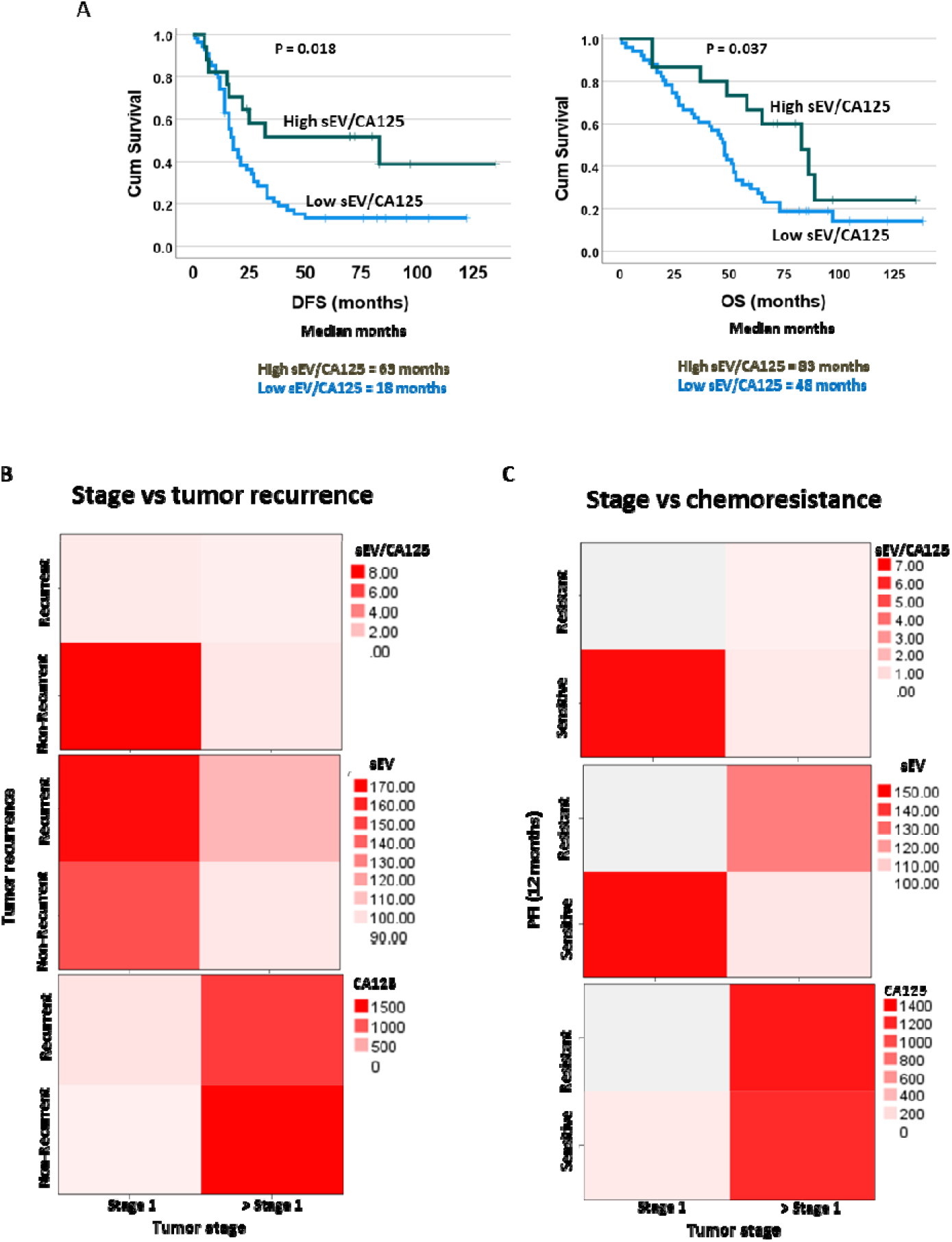
High levels of sEV/CA125 provide favorable survival outcomes to OVCA patients. sEV/CA125 levels in OVCA patients (N=99) were correlated with progression-free survival (PFS) and overall survival (OS). Kaplan-Meier survival curves with dichotomized sEV/CA125 levels (low and high groups, cut-off = 68.8) and log rank test were used to compare the survival distributions between the groups. N = number of patients (B) sEV/CA125 predicts more than one clinical outcome of OVCA. SEV/CA125, sEV and CA125 were used in a heat map analyses to predict stage 1 and tumor recurrence as well as stage 1 and chemoresistance.

Kaplan-Meier survival curves with dichotomized sEV/CA125 ratio (low vs. high, cut-off = 68.8) and log rank test were used to compare the survival distributions between the groups (Fig. 6A). We observed that patients with high sEV/CA125 had significantly improved disease free survival (DFS; 63 months vs. 18 months, p=0.018) as well as overall survival (OS; 83 months vs. 48 months, p=0.037) compared with patients with low sEV/CA125 (Fig. 6A). We further investigated if sEV/CA125 could be used to predict multiple clinical outcomes using a heat map (Fig. 6B-C). Compared with sEV and CA125 individually, sEV/CA125 ratio provided a significant prediction value for multiple clinical outcomes. Patients with increased sEV/CA125 were mostly in stage 1 and did not develop recurrence or resistance to treatment (Fig. 6B and C). SEV and CA125 concentrations alone were not predictive of clinical outcomes.

## DISCUSSION

For the first time, we have shown evidence that pGSN plays a regulatory role in sEV secretion as well as sEV-mediated release of CDDP; factors that are involved in OVCA chemoresistance. Additionally, we extended the application of our novel sEV-based biosensor to a small OVCA patient cohort to develop a highly sensitive diagnostic platform for the detection of early stage OVCA and prediction of chemoresponsiveness.

Drug efflux is a key determinant of chemoresistance in a lot cancer types, including OVCA cancer (34-37). Most studies have identified multi-drug resistant proteins such as p-glycoprotein to be responsible for drug efflux in cancer cells, thus rendering chemotherapeutic agents unable to induce death in cancer cells (36, 37). In this present study, we have demonstrated that sEVs serve as a vehicle for exporting chemotherapeutic agents from OVCA cancer cells rather than using drug transport channels. Chemoresistant OVCA cells treated with CDDP secreted increased amount of sEVs as well as sEV containing CDDP compared with their sensitive counterparts. This could potentially explain why targeting drug transport channels such as P-glycoprotein has not yielded any therapeutic benefit in OVCA given the sEVs role in drug efflux. This suggests that sEV secretion is a potential therapeutic target to tackle chemoresistance in OVCA.

Targeting sEV secretion also demands understating the mechanism underlying why chemoresistant cells secrete more sEVs and sEV-CDDP. pGSN is overexpressed in chemoresistant OVCA compared with chemosensitive cells and transported via sEVs (10). The positive association between pGSN and sEVs made us investigate the role of pGSN in sEV secretion. We observed that silencing pGSN reduced the expression of CTTN (involved is sEV release mechanisms) in chemoresistant cells resulting in decreased sEV secretion and EV-CDDP release. The opposite happened when pGSN was overexpressed in chemosensitive OVCA cells. Targeting pGSN in the chemoresistant cells resulted in increased accumulation of CDDP leading to increased apoptosis, suggesting that pGSN is a key regulator of CTTN expression. These findings are consistent with the study in which pGSN and CTTN were found to be upregulated and co-localized in resistant pancreatic cancer cell lines. Furthermore, recent studies have shown that proteins rich in sulphur and amines have high affinity for cisplatin binding. Since pGSN has these properties as well as more than 10 metal binding sites, there is a possibility that it could directly bind to CDDP and facilitate their packaging into sEVs for secretion. This will be worth investigating in the future.

Late diagnosis is a huge challenge to therapeutic success and patient survival. Developing a highly sensitive diagnostic platform for early OVCA diagnosis is of urgent need. With heterogeneity and protein interference as major challenges when using SERS for sEV analysis, we have developed a biosensor to overcome this challenge. This biosensor is able to react with sEVs and CDDP simultaneously; a strategy that enables us to quantitate both sEVs and CDDP simultaneously using SERS. This strategy is highly sensitive and superior to other studies that have developed gold nanoparticles for sEV analysis alone.

We examined the diagnostic utility of our sEV-based biosensor in a small pre-operative OVCA cohort. Although sEVs could be detected and quantified, their diagnostic value was not as strong as when combined with CA125. This could potentially be due to the fact that the biosensor recognized sEVs from origins other than OVCA. To overcome this, we used a ratio of sEVs to CA125. We observed that the sEV/CA125 ratio had a significantly improved diagnostic value. sEV/CA125 was significantly elevated in stage 1 patients compared to stages 2-4 and outperformed CA125 and sEV in identifying stage 1 disease. This is a promising marker given conventional biomarkers have only provided modest diagnostic value. Interestingly, sEV/CA125 also outperformed CA125 and sEVs in predicting tumor recurrence, chemoresistance, residual disease and patient survival. This is exciting given there is no reliable biomarker to predict tumor recurrence and chemoresistance. OVCA patients with increased sEV/CA125 had prolonged survival compared with those with low sEV/CA125, suggesting that sEV/CA125 could be used as a potential marker for early stage OVCA and predict patient survival outcomes prior treatment or surgery. With limited biomarker options for early stage OVCA and prediction of chemoresistance, sEV/CA125 could be further investigated to provide the needed diagnostic tool.

## CONCLUSION

We have demonstrated that pGSN regulates sEV and sEV-CDDP secretions via CTTN upregulation leading to decreased intracellular CDDP accumulation and apoptosis. These processes further suppress chemo-responsiveness in OVCA cells. Additionally, we have applied a novel biosensor to develop a platform that is effective in identifying stage 1 disease and predicting tumor recurrence and chemoresistance. Although these findings are promising and have an important clinical relevance, further validation of the platform is needed in a larger patient cohort and animal models.

## Data Availability

All data produced in the present study are available upon reasonable request to the authors

