## Supplementary Figures for "The application of an extracellular vesicle-based biosensor in early diagnosis and prediction of chemoresponsiveness in ovarian cancer"

### Slide 1
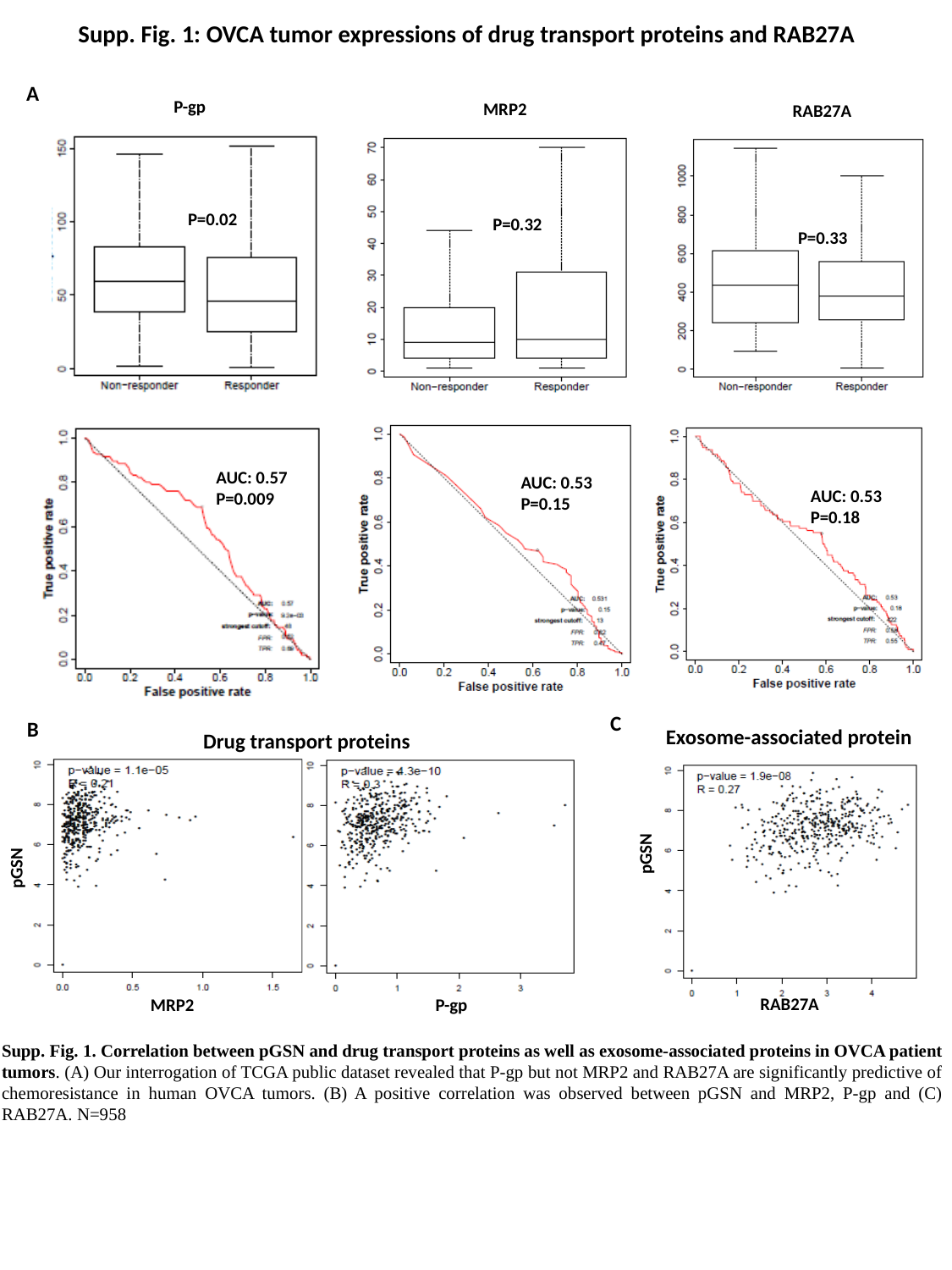

Supp. Fig. 1: OVCA tumor expressions of drug transport proteins and RAB27A
A
P-gp
P=0.02
AUC: 0.57
P=0.009
MRP2
P=0.32
AUC: 0.53
P=0.15
RAB27A
P=0.33
AUC: 0.53
P=0.18
C
B
Exosome-associated protein
Drug transport proteins
pGSN
pGSN
RAB27A
MRP2
P-gp
Supp. Fig. 1. Correlation between pGSN and drug transport proteins as well as exosome-associated proteins in OVCA patient tumors. (A) Our interrogation of TCGA public dataset revealed that P-gp but not MRP2 and RAB27A are significantly predictive of chemoresistance in human OVCA tumors. (B) A positive correlation was observed between pGSN and MRP2, P-gp and (C) RAB27A. N=958

### Slide 2
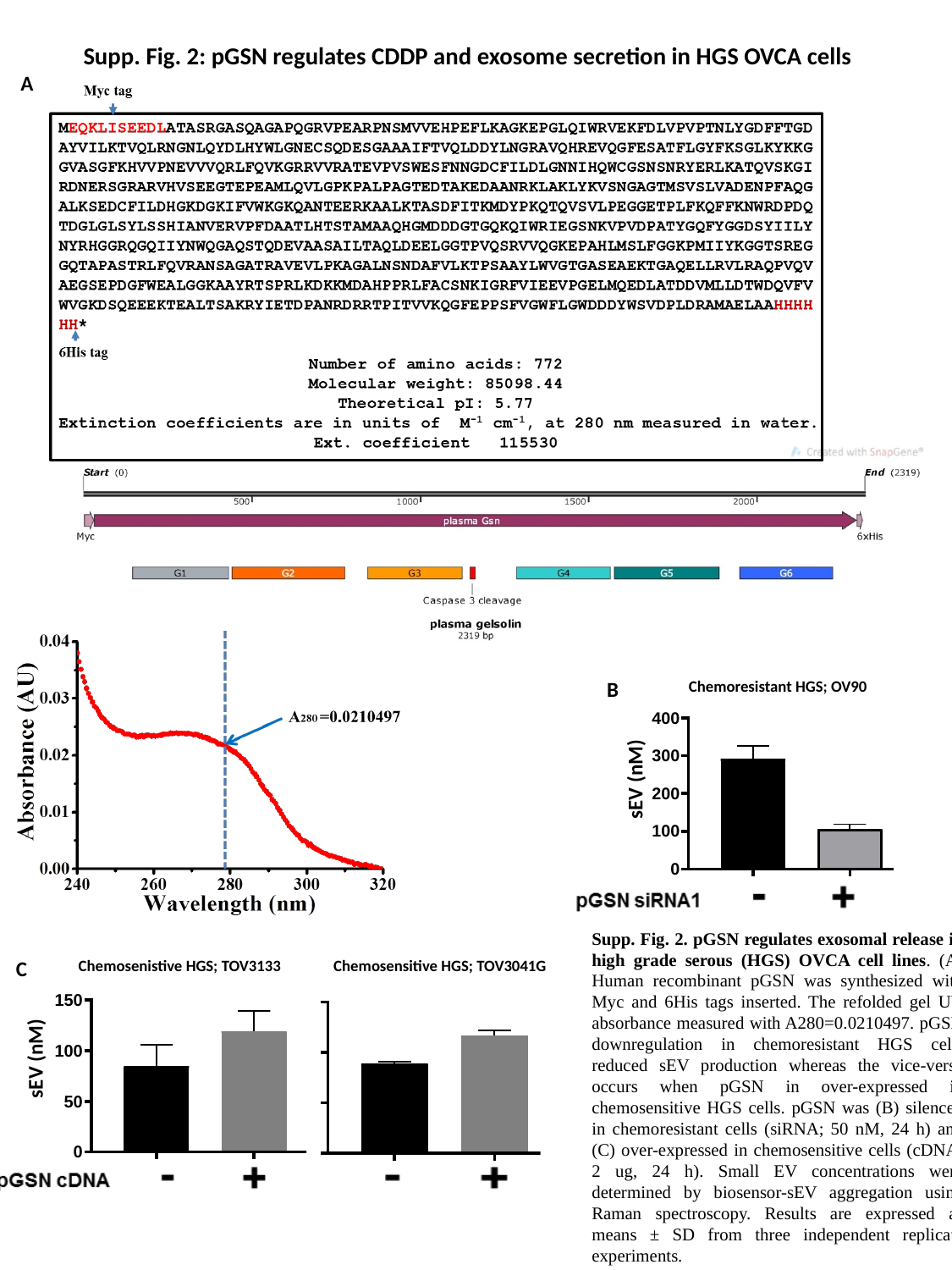

Supp. Fig. 2: pGSN regulates CDDP and exosome secretion in HGS OVCA cells
A
Chemoresistant HGS; OV90
B
sEV (nM)
Supp. Fig. 2. pGSN regulates exosomal release in high grade serous (HGS) OVCA cell lines. (A) Human recombinant pGSN was synthesized with Myc and 6His tags inserted. The refolded gel UV absorbance measured with A280=0.0210497. pGSN downregulation in chemoresistant HGS cells reduced sEV production whereas the vice-versa occurs when pGSN in over-expressed in chemosensitive HGS cells. pGSN was (B) silenced in chemoresistant cells (siRNA; 50 nM, 24 h) and (C) over-expressed in chemosensitive cells (cDNA; 2 ug, 24 h). Small EV concentrations were determined by biosensor-sEV aggregation using Raman spectroscopy. Results are expressed as means ± SD from three independent replicate experiments.
C
Chemosenistive HGS; TOV3133
Chemosensitive HGS; TOV3041G
sEV (nM)

### Slide 3
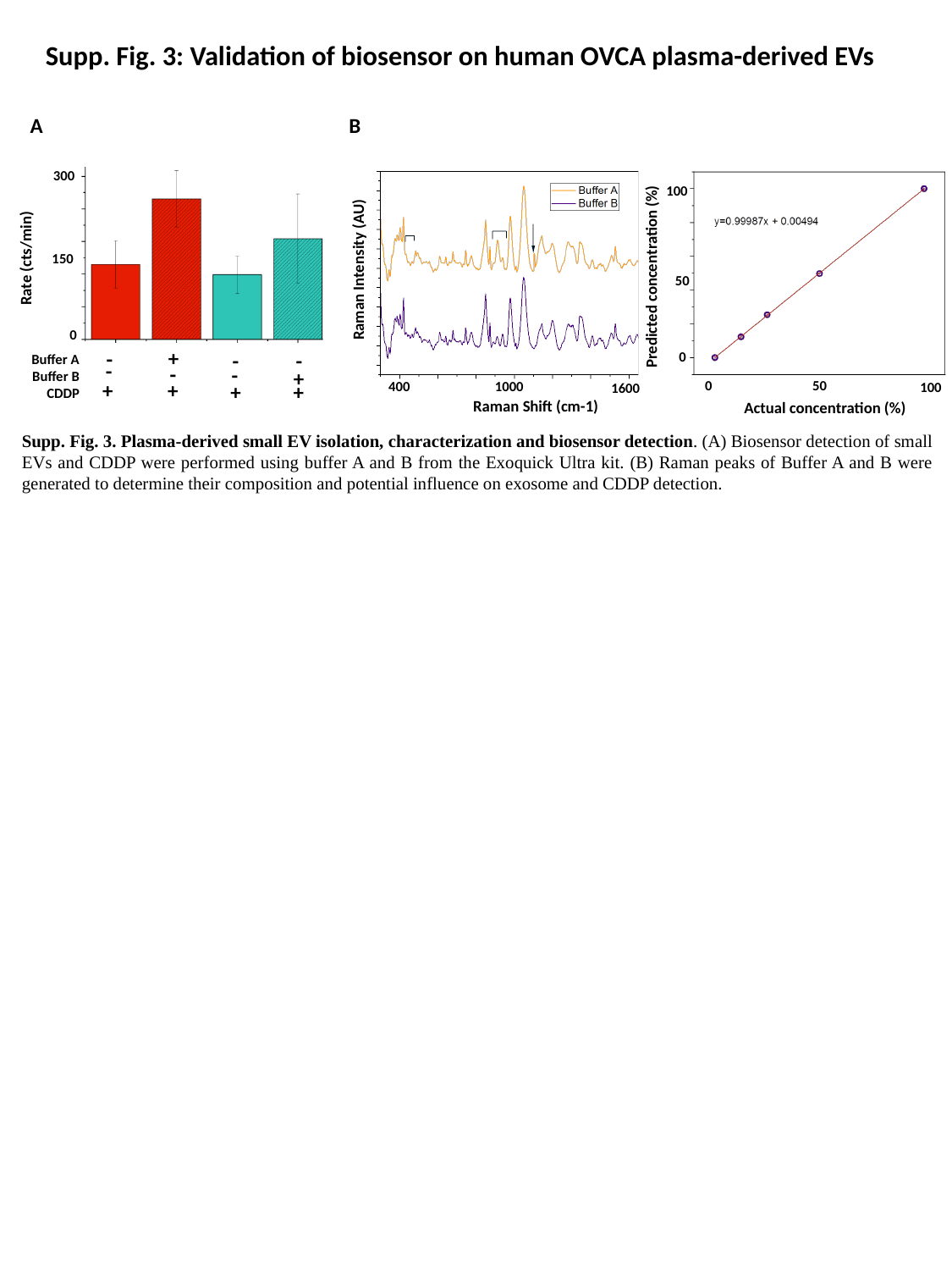

Supp. Fig. 3: Validation of biosensor on human OVCA plasma-derived EVs
A
B
300
Rate (cts/min)
150
0
+
-
-
-
Buffer A
Buffer B
CDDP
-
-
-
+
+
+
+
+
100
Raman Intensity (AU)
Predicted concentration (%)
50
0
50
0
1000
400
100
1600
Raman Shift (cm-1)
Actual concentration (%)
Supp. Fig. 3. Plasma-derived small EV isolation, characterization and biosensor detection. (A) Biosensor detection of small EVs and CDDP were performed using buffer A and B from the Exoquick Ultra kit. (B) Raman peaks of Buffer A and B were generated to determine their composition and potential influence on exosome and CDDP detection.

### Slide 4
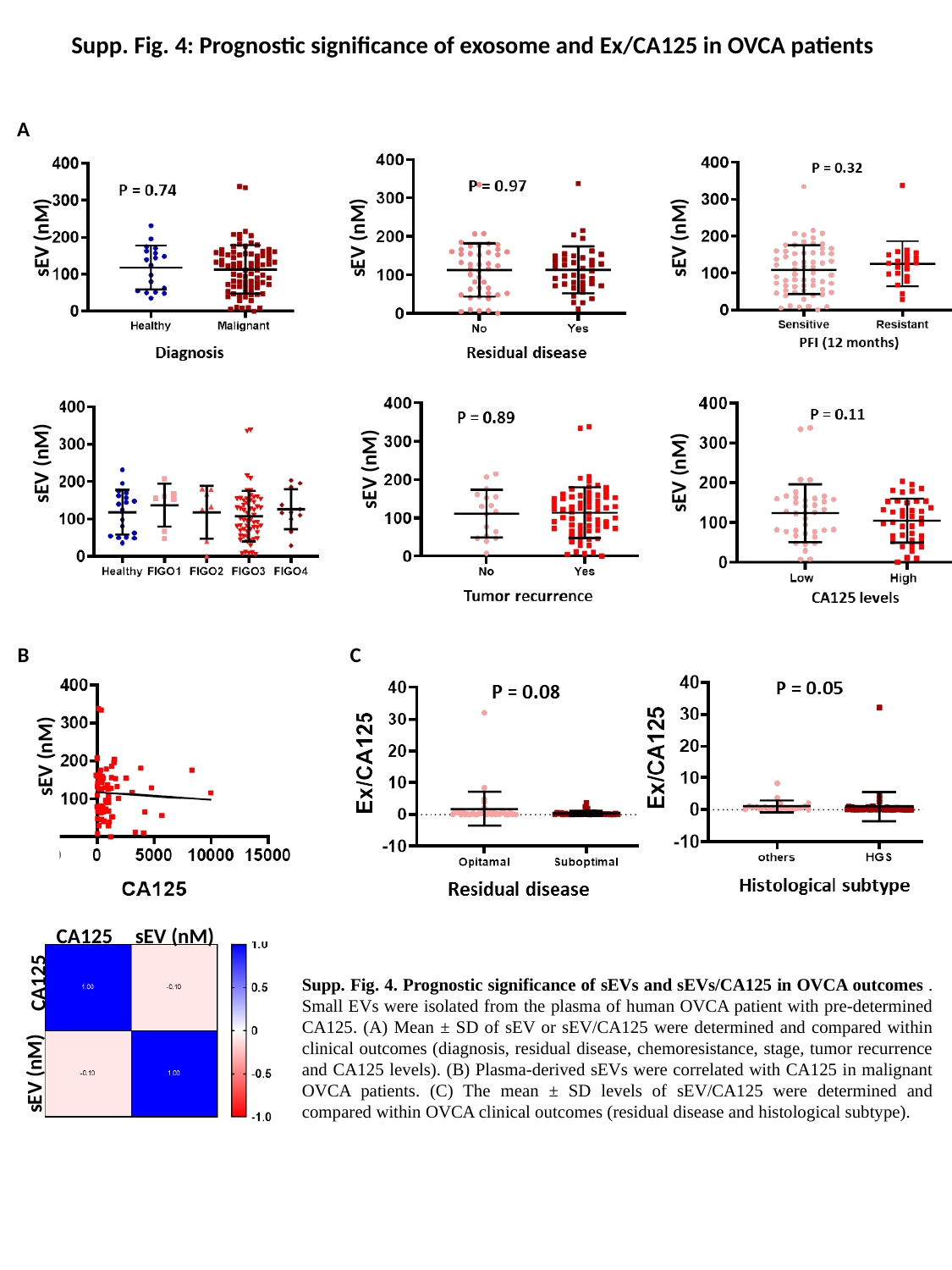

Supp. Fig. 4: Prognostic significance of exosome and Ex/CA125 in OVCA patients
A
sEV (nM)
sEV (nM)
sEV (nM)
sEV (nM)
sEV (nM)
sEV (nM)
B
C
sEV (nM)
CA125
sEV (nM)
CA125
sEV (nM)
Supp. Fig. 4. Prognostic significance of sEVs and sEVs/CA125 in OVCA outcomes . Small EVs were isolated from the plasma of human OVCA patient with pre-determined CA125. (A) Mean ± SD of sEV or sEV/CA125 were determined and compared within clinical outcomes (diagnosis, residual disease, chemoresistance, stage, tumor recurrence and CA125 levels). (B) Plasma-derived sEVs were correlated with CA125 in malignant OVCA patients. (C) The mean ± SD levels of sEV/CA125 were determined and compared within OVCA clinical outcomes (residual disease and histological subtype).
