## Supplementary Tables for "The application of an extracellular vesicle-based biosensor in early diagnosis and prediction of chemoresponsiveness in ovarian cancer"

**Running Title: Extracellular vesicle-mediated CDDP secretion in OVCA chemoresistance**

*Correspondance:

Dr. Benjamin K Tsang, Chronic Disease Program, Ottawa Hospital Research Institute, The Ottawa Hospital (General Campus), Ottawa, Canada K1H 8L6; Tel: 1-613-798-5555 ext 72926;

**Supplementary Table 1. Characteristics of patients and subjects**

| Variable | Number of Patients |
| --- | --- |
| Age (Range; 36 – 82 years)  ≤61  >61 | 51  48 |
| Stage (FIGO)  1  2  3  4 | 10  11  67  11 |
| Histological Subtypes  Not verified  High Grade Serous (HGS)  Low Grade Serous (LGS) | 26  69  4 |
| Residual disease (RD)  ≤1 cm  >1 cm | 50  42 |
| Healthy Subjects | 20 |

FIGO, International Federation of Gynecology and Obstetrics.

**Supplementary Table 2. Information on antibodies and reagents**

| Primary Antibodies | | | | | | Secondary Antibodies | | | | | |
| --- | --- | --- | --- | --- | --- | --- | --- | --- | --- | --- | --- |
| Application | **Target** | **Antibody/reagent** | **Company** | **Catalog #** | **Dilution** | **Antibody** | **Conjugate** | **Company** | **Catalog #** | **Dilution** | **Note** |
| WB | pGSN | Anti-pGSN Goat polyclonal | Antibodies online(Atlanta,USA) | ABIN1019662 | 1:1000 | Dnk polyclonal to Goat IgG | HRP | Abcam (Toronto, Canada) | Ab97110 | 1:2000 |  |
| WB | GAPDH | Anti-actin mouse monoclonal | Abcam (Toronto, Canada) | ab8226 | 1:1000 | Goat Anti-mouse IgG (H+L) | HRP | Bio-Rad (Mississauga, Canada) | 170-6516 | 1:2000 |  |
| WB | CD9 |  |  |  |  |  |  |  |  |  |  |
| WB | CD63 |  |  |  |  |  |  |  |  |  |  |
| WB | CD81 |  |  |  |  |  |  |  |  |  |  |
| WB | GM130 |  |  |  |  |  |  |  |  |  |  |
| WB | P-gp |  |  |  |  |  |  |  |  |  |  |
| WB | CTTN |  |  |  |  |  |  |  |  |  |  |
| WB | RAB27A |  |  |  |  |  |  |  |  |  |  |
| IF | CD63 | pCT-CD63-GFP |  |  |  |  |  |  |  |  |  |
|  | Nuclei | Dapi |  |  |  |  |  |  |  |  |  |
|  | EVs | Exoquick Ultra |  |  |  |  |  |  |  |  |  |

| Cell line | Tumor origin | TP53 status | Other | Chemosensitivity |
| --- | --- | --- | --- | --- |
| A2780s | Ovarian endometroid adenocarcinoma | Wild-type | PTEN/ARID1A | Sensitive |
| A2780cp | Ovarian endometroid adenocarcinoma | Mutant  V127F, R260S | PTEN/ARID1A | Resistant |
| OV90 | High grade serous ovarian cancer | Mutant  Ser215Arg | None Detected | Resistant |
| TOV3133G | Serous-papillary adenocarcinoma | Nonsense | None Detected | Sensitive |
| TOV3041G | Serous adenocarcinoma | Wild-type | None Detected | Sensitive |

**Supplementary Table 3. Information on OVCA cell lines**

**Information on OVCA cell lines**: The characterization of these cell lines have been verified in previous literature (Anglesio et al.,2013; Leroy et al., 2014; Provencher et al., 2000; Fleury et al., 2015; Letourneau et al., 2012; Ouellet et al., 2008; ).

**Supplementary Table 4: Customized siRNA oligonucleotide duplexes**

| **Product** | **Target** | **Species** | **Company** | **Catalog #** | **Target sequence** | **Anti-sense sequence** | **Position on mRNA** |
| --- | --- | --- | --- | --- | --- | --- | --- |
| siRNA 1 | pGSN | Human | IDT (Iowa, USA) | N/A | GCGACCCGAGGCCGCGGCU | AGCCGCGGCCUCGGGUCGC | 12 |
| siRNA 2 | pGSN | Human | IDT (Iowa, USA) | N/A | UGCCCGAGGCGCGGCCCAA | UUGGGCCGCGCCUCGGGCA | 192 |

IRS, immunoreactive score; pGSN, plasma gelsolin
